# Unveiling the burden: Depression and its determinants among Bangladeshi medical students - insights from the MINI diagnostic tool

**DOI:** 10.1101/2025.01.30.25321378

**Authors:** Zarin Tasnim Maliha, Sayeda Nazmun Nahar, Dipak Kumar Mitra, Nadira Sultana Kakoly, Kamrun Nahar Koly, Md Humayun Kabir Talukder, Helal Uddin Ahmed, Rajat Das Gupta, M. Tasdik Hasan

## Abstract

Mental health challenges are widespread among medical students worldwide, exacerbated by a lack of help-seeking behavior within this population. In resource-constrained healthcare systems like Bangladesh, medical students face additional barriers to expressing their struggles or seeking support. This study is the first in Bangladesh to use the MINI, a confirmatory clinical diagnostic tool, to assess the prevalence of depression among medical students. This cross-sectional study employed a convenient sampling technique, with 529 students from across Bangladesh participating via online surveys. The survey included questions on relevant sociodemographic factors and the PHQ-9 depression screening tool. Subsequently, depression was confirmed through clinical diagnosis using the MINI, conducted via online interviews over Zoom and phone calls. The study was carried out from June 2020 to February 2021, and data analysis was performed using SPSS software (version 22.0). Among the medical students surveyed using PHQ-9, 31.5% exhibited moderate levels of depression, 38.7% showed mild depression, and 29.8% reported minimal depression. Clinical diagnoses conducted using the MINI tool confirmed depression in 50 PHQ-9-positive cases. Of these, 17 (34%) were categorized as having “no depression,” 27 (54%) were diagnosed with recent depression, 16 (32%) had a history of past depression, and 15 (30%) experienced recurrent depression. Depression was more prevalent among female medical students, particularly those living away from their families from the start of their degree. First-year students were found to have the strongest association with depression. This study reported that one-third of Bangladeshi medical students experienced moderate to severe depression. The findings underscore the need for targeted psychosocial interventions and further exploration of socio-demographic factors. These results aim to guide researchers and policymakers in addressing the mental health needs of this population through effective support systems and surveillance frameworks.

## Introduction

Globally, medical schools are responsible for providing adequate knowledge to the students throughout the medical years to produce competent doctors in the future. And they also make sure the medical curriculum is outlined as a full package of quality study training material [1] though it’s hard for all medical students to adapt with such study pressure. A degree of stress is required to motivate them throughout the difficult medical years though pressure and stress lead them to develop emotional and mental distress. Frustrations, depression symptoms, anxiety are the common syndrome among them. Also, they share virtually no focus when it comes to monitoring their physical health let alone their mental health. The desire for absolute perfection and the great expectations of those around them push young medical students to the edge of their mental stability. Stress, anxiety, depressive symptoms, and panic attacks are becoming integral parts of medical students. These mental health problems are indicative of clinical depression, frustration, OCD, other mental disorders, and sometimes suicidal attitudes.

Many studies have reflected a higher relative susceptibility of medical students to stress and anxiety levels than students pursuing degrees at regular universities [2]. A medical student begins to accumulate mental problems right from the start of college, where he has to go through a packed schedule of classes, assessments, lab work, and professional exams, etc. Most students cannot keep up with the pace of medical life and thus begins to sink in constant stress and anxiety [5].

The number of students trying to escape mental illness has skyrocketed. According to a contemporary study of medical school students in China, it was found that approximately 29%, 21%, and 11% of respondents suffered from depression, anxiety and suicidal ideation [3]. The British Medical Association reports that over a period of one and a half years, in 2018 six medical students committed suicide [4]. In a study of a medical school in the US, it was found that 25% of students suffered from moderate to severe anxiety and 9% of students suffered from depression [9]. Depression is a common mental disorder, characterized by sadness, loss of interest or pleasure, feelings of guilt or low self-worth, disturbed sleep or appetite, feelings of tiredness and poor concentration. In a study of African first-year medical students, it was found that 64.5% of respondents were mentally affected by various episodes of stress and depression [7].

Bangladesh ranks first among the countries with the most medical students suffering from mental disorders. Due to 6 years of university study (5 years of formal study and 1 year of internship), medical students often experience mental health problems [8]. In a cross-sectional study conducted in Dhaka, Bangladesh, it was found that 38.9% of students suffered from depression and 17.6% of them had had suicidal thoughts at least once in their lives [5]. The same study concluded that 33.5% of medical students suffer from mental health problems while 16.8% of the adult population of Bangladesh is in a state of mental instability, according to the report of the National Mental Health Survey 2018/19. It shows that the likelihood of mental health problems in medical students is almost twice that of the average adult.

So, it’s fair to say Bangladesh has entered a worrying era where many medical students struggle with depression, anxiety, stress and other mental health related issues. If things are not investigated properly, students with various mental health problems are at risk of failing their professional careers and above all, they may even face serious consequences in their personal lives as a result of mental health negligence. story. In this report, quantitative results on the prevalence of depressive symptoms, clinical depression, and associated factors are being presented.

14% of medical students suffered from mild to moderate depression worldwide. [19] A psychiatrist is assigned to 7,500 people in the United States and Bangladesh, the scenario is even worse. Only 0.4% work as a psychologist here. A psychiatrist is available for millions of people in Bangladesh. [20] There is a gap in the provision of mental health services as well as in mental health screening for medical students in Bangladesh. This study could provide a way to identify gaps in health systems by assessing their real situation. The aim was to have a clear picture of the reality of mental health issues with a particular focus on depression among medical students in Bangladesh. This survey will present a real-life situation, helping to create an opportunity to identify challenges and gaps. The objective of this study was to recognize the main areas of support required by medical students of different economies, ethnicities and genders. It is expected that this survey will generate ideas for creating opportunities to address mental health through on-campus programs / interventions that enhance lifelong learning, education, including government and private medical colleges.

Studying medicine is a challenging endeavor worldwide, including in resource poor settings like Bangladesh. Medical students are often expected to work harder than peers in other academic disciplines, requiring exceptional mental toughness, endurance, and intelligence [3]. Undergraduate medical students face a particularly stressful period of training, characterized by intense mental and physical demands compared to other fields of study [1]. Consequently, they encounter numerous mental and emotional challenges during their medical education. The rigorous journey of becoming a doctor often predisposes students to mental health issues, including depression, anxiety, and other psychological disorders. Young age further increases their vulnerability to these challenges [3].

Globally, medical students exhibit a higher prevalence of mental health disorders than students in other fields, and this trend is on the rise. For instance, one study reported that 27.2% of medical students experienced depression or depressive symptoms, while 11% had suicidal ideation [17]. While a certain level of stress can motivate students, excessive stress combined with emotional distress often predisposes them to severe mental health issues. Depression, in particular, is the most common mental health disorder among medical students, primarily due to the overwhelming academic and psychological pressures they face [6].

In Bangladesh, undergraduate medical students must study for at least five years, followed by a one-year internship, to earn their medical degree [23]. This prolonged and intense academic journey significantly contributes to mental health challenges [35]. Many students develop anxiety and depression due to extreme academic pressure during these years. Socioeconomic and cultural factors further exacerbate these issues, as students struggle to meet societal expectations while managing strenuous schedules. Such challenges often lead to mental health disorders, with some students even contemplating suicide. The suicide rate among Bangladeshi medical students has been increasing, with several cases reported in recent years [24].

Currently, Bangladesh has approximately 133 registered medical schools, including public and private institutions [23]. Thousands of students endure the demanding process of becoming physicians, but not all are equipped to handle the pressure. Many struggles emotionally and mentally, leading to mental health disorders. A study of 399 medical students in Bangladesh revealed that 39.1% experienced depression, and 18.8% reported suicidal ideation following prolonged depressive episodes, which is higher than in many other countries [2]. The rigorous academic demands, combined with the lack of a supportive learning environment, contribute significantly to mental health issues among Bangladeshi medical students. Another recent study found that 54% of medical students experienced high levels of stress, based on data collected from 990 students across two public and six private medical schools [1]. Unfortunately, despite the high prevalence of mental health issues, medical students are less likely to seek professional help. Barriers include rigid schedules, stigma surrounding mental health, and fear of being perceived as unfit for the profession [25]. This reluctance exacerbates their psychological distress, leading to long-term consequences such as chronic depression and burnout [9]. Cultural factors also play a significant role in preventing medical students from seeking help. Deep-rooted beliefs in prioritizing patient care over personal well-being discourage students from addressing their mental health needs. Fear of stigma and confidentiality breaches further deters them from consulting mental health professionals. As a result, many students suffer in silence, often leading to academic dropout or severe mental health crises [26].

Although several studies have highlighted the alarming prevalence of mental health issues among Bangladeshi medical students, driven by academic, social, and cultural factors, none to date have reported confirmatory diagnosis of depression using a standardized diagnostic tool [5, 11]. The primary objective of this study was to estimate the burden of depression among medical students in Bangladesh using the MINI diagnostic tool, a recognized confirmatory clinical diagnostic instrument. Specifically, the study aimed to assess the prevalence of depression in this population and identify the associated factors contributing to its occurrence.

## Methods

### Study Design and Setting

This cross-sectional study investigated depression among medical students in Bangladesh. The study was conducted from June 2020 to February 2021, with data collected online from students enrolled in 28 public and 29 private medical colleges nationwide. Data collection period was from 22nd January, 2021 to 12th February, 2021. Participants included Bangladeshi students currently pursuing MBBS and BDS degrees in their 1st to 5th year of study. Students of non-Bangladeshi nationality and those diagnosed with chronic diseases, chronic depression, or known substance abuse were excluded from the study.

### Sampling Method and Sample Size

A convenience sampling method was used to recruit participants. The initial sample size was 384, based on standard sampling calculations. However, due to the shift from face-to-face data collection to an online format during the COVID-19 pandemic, oversampling was applied.

A total of 600 medical students were approached, resulting in 529 valid responses, ensuring adequate representation and minimizing response bias.

### Instruments Used

- Screening for Depression: The Patient Health Questionnaire-9 (PHQ-9) was used to screen for depressive symptoms.
- Diagnosis Confirmation: The Mini International Neuropsychiatric Interview (M.I.N.I.), a brief structured diagnostic interview based on DSM-III-R, DSM-IV, DSM-5, and ICD-10 criteria, was used to confirm depression.

A subset of 50 participants, identified as depressed through the PHQ-9 scale, were randomly selected for diagnostic confirmation using the M.I.N.I.

### Data Collection

Data were collected via an online survey using a structured questionnaire distributed through institutional networks, email, and social media platforms. The online approach ensured participant safety and compliance with COVID-19 restrictions while facilitating broader geographic reach.

### Data Management and Analysis

Through descriptive statistics, depressive symptoms, clinical depression and associated factors of depression were presented in this report. In descriptive analysis, both central tendency (i.e., percentage, mean and frequency) and dispersion statistic (range, standard deviation) were calculated for participant demographic characteristics among different study groups. For nominal/categorical variables, chi-square test and for continuous variable, two sample independent t-tests were performed to see the observed relationship with study groups. P-value<0.05 (two-tailed) was considered the margin of statistical significance for all tests. All the data were entered carefully, managed, cleaned by the study team and analyzed using SPSS software 22.0 version.

### Quality Control and Assurance

Periodic assessments of data quality and timeliness, participant recruitment and other factors that can affect study outcome were periodically assessed by the lead researcher. For clinically diagnosed depression cases we provided contact information of National Institute of Mental Health, Dhaka as the primary contact. No follows up was considered due to resource constraints.

### Ethical Considerations

Ethical approval (IRB Reference no: 2020/OR-NSU/IRB-No.0502) was obtained from North South University-Bangladesh. Electronic informed consent was taken from the participants while collecting data online. Privacy & confidentiality of the respondents during data collection were maintained strictly.

## Results

A total of 529 questionnaires met the inclusion criteria for this study. The mean age (± standard deviation, SD) of the participants was 22.75 years (± 1.98), with the majority, 430 (81.6%), belonging to the late adolescence age group (18–24 years). Regarding gender, most participants were female, comprising 310 (58.8%) of the sample, while 5 (0.9%) participants chose not to disclose their gender.

Among students enrolled in the five-year MBBS program, the majority were in their 3rd year, totaling 322 (61.1%), followed by those in their 5th year (22.8%) and 4th year (15.7%). A significant proportion of participants, 322 (61.1%), lived in hostels, while 180 (34.2%) resided with their families, and the remainder stayed in shared accommodations such as messes or other living arrangements.

Among lifestyle habits, smoking was the most prevalent, reported by 95 (18%) participants, followed by caffeine consumption (59, 11.2%), alcohol use (26, 4.9%), and substance abuse (18, 3.4%). The majority of participants came from well-educated families, with 82.9% of fathers and 74% of mothers having completed higher secondary education. Furthermore, 44.4% of fathers and 28.8% of mothers had achieved advanced academic qualifications, such as postgraduate degrees.

Regarding family size, 215 (40.8%) participants belonged to nuclear families (four or fewer members), while the majority, 296 (56.2%), had families with 5–8 members. Additionally, more than 182 (34%) participants reported having elderly family members within their households.

The economic status of participants’ families was comparatively better than the national average. Approximately 35% of participants reported a monthly family income between 25,001 and 50,000 Bangladeshi Taka (BDT), about 30% reported income between 50,001 and 99,999 BDT, and over 15% reported a family income of 100,000 BDT or more. Regarding financial independence, 357 (67.7%) participants relied on various sources for pocket money, while only a few, 105 (19.9%), had a personal income source. Consequently, only 65 (12.3%) of participants could contribute financially to their families (Table 1).

**Table 1.** Sociodemographic characteristics of the study participants.

| Variables with Categories | Frequency (No.) | Percent (%) |
| --- | --- | --- |
| **Age Group** |  |  |
| 18-24 years | 430 | 81.6 |
| 25-29 years | 97 | 18.4 |
| **Gender** |  |  |
| Female | 310 | 58.8 |
| Male | 212 | 40.2 |
| Not disclosed | 5 | 0.9 |
| **Year of education** |  |  |
| 1<sup>st</sup> year medical student | 70 | 13.3 |
| 2<sup>nd</sup> year medical student | 52 | 9.9 |
| 3<sup>rd</sup> year medical student | 202 | 38.3 |
| 4<sup>th</sup> year medical student | 83 | 15.7 |
| 5<sup>th</sup> year medical student | 120 | 22.8 |
| **Living Place** |  |  |
| Hostel | 322 | 61.1 |
| Mess | 18 | 3.4 |
| With Family | 180 | 34.2 |
| Others | 7 | 1.3 |
| **Common habits** |  |  |
| Smoking | 95 | 18 |
| Alcoholism | 26 | 4.9 |
| Caffeine | 59 | 11.2 |
| Substance abuse | 18 | 3.4 |
| **Father's Education** |  |  |
| Primary | 11 | 2.1 |
| Secondary | 55 | 10.4 |
| Higher Secondary | 109 | 20.7 |
| Undergraduate | 94 | 17.8 |
| Post-graduate | 234 | 44.4 |
| Others | 24 | 4.6 |
| **Mother's Education** |  |  |
| Primary | 18 | 3.4 |
| Secondary | 99 | 18.8 |
| Higher Secondary | 134 | 25.5 |
| Undergraduate | 104 | 19.7 |
| Post-graduate | 152 | 28.8 |
| Others | 20 | 3.8 |
| **Number of members in family** |  |  |
| ≤ 4 members | 215 | 40.8 |
| 5-8 members | 296 | 56.2 |
| ≥ 9 members | 16 | 3.0 |
| **Presence of aged family members in family** |  |  |
| Yes | 182 | 34.5 |
| No | 345 | 65.5 |
| **Monthly total family income (BDT)** |  |  |
| ≤ 25000 | 109 | 20.7 |
| 25001-50000 | 183 | 34.7 |
| 50001-99999 | 155 | 29.4 |
| ≥ 100000 | 80 | 15.2 |
| **Personal income** |  |  |
| Yes | 105 | 19.9 |
| No | 422 | 80.1 |
| **Family contribution** |  |  |
| Yes | 65 | 12.3 |
| No | 462 | 87.7 |
| **Get pocket money from various source** |  |  |
| Yes | 357 | 67.7 |
| No | 170 | 32.3 |

### Prevalence of depression among the participants

According to the study findings, 4.6% of study participants had severe depression, 11% had moderately severe depression, 15.9% had moderate depression, 38.7% had mild depression, and 29.8% had minimal depression (Fig 1).

**Fig 1.**
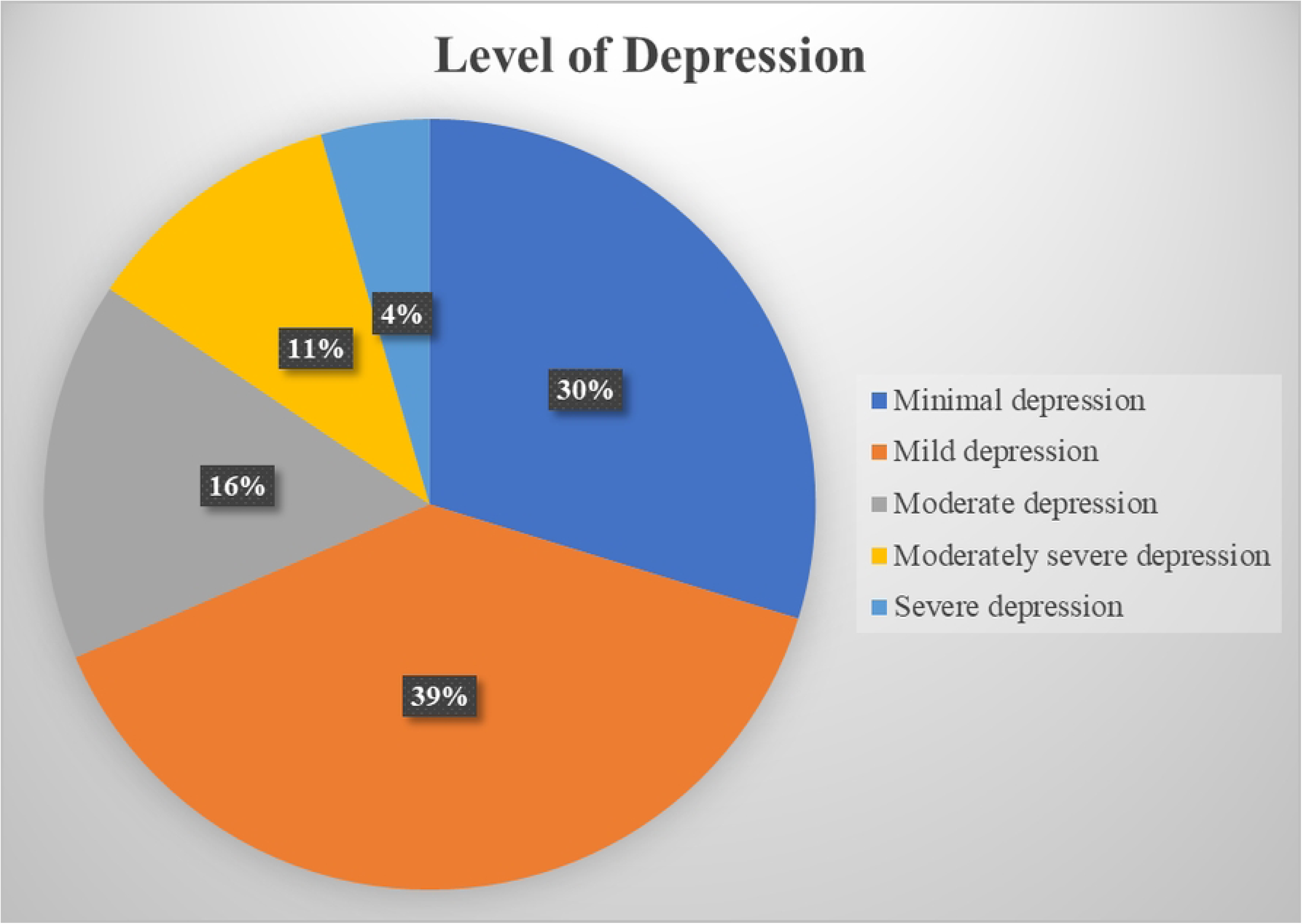
Depression symptoms of the participants according to PHQ 9 scale. Distribution of depression levels among medical students: minimal, mild, moderate, moderately severe, and severe.

### Depression confirmation by the M.I.N.I psychiatric diagnostic instrument

In MINI assessment depression is categorized into ‘No’, ‘Recent’, ‘Past’, and ‘Recurrent’. Among the participants of MINI 17 (34%) was found under the ‘No depression’ category though all of them were found depressed in the PHQ-9 scale. Moreover, 27 (54%) of the participants had recent depression, 16 (32%) had past depression, and 15 (30%) of the participants had recurrent depression (Fig 2).

**Fig 2.**
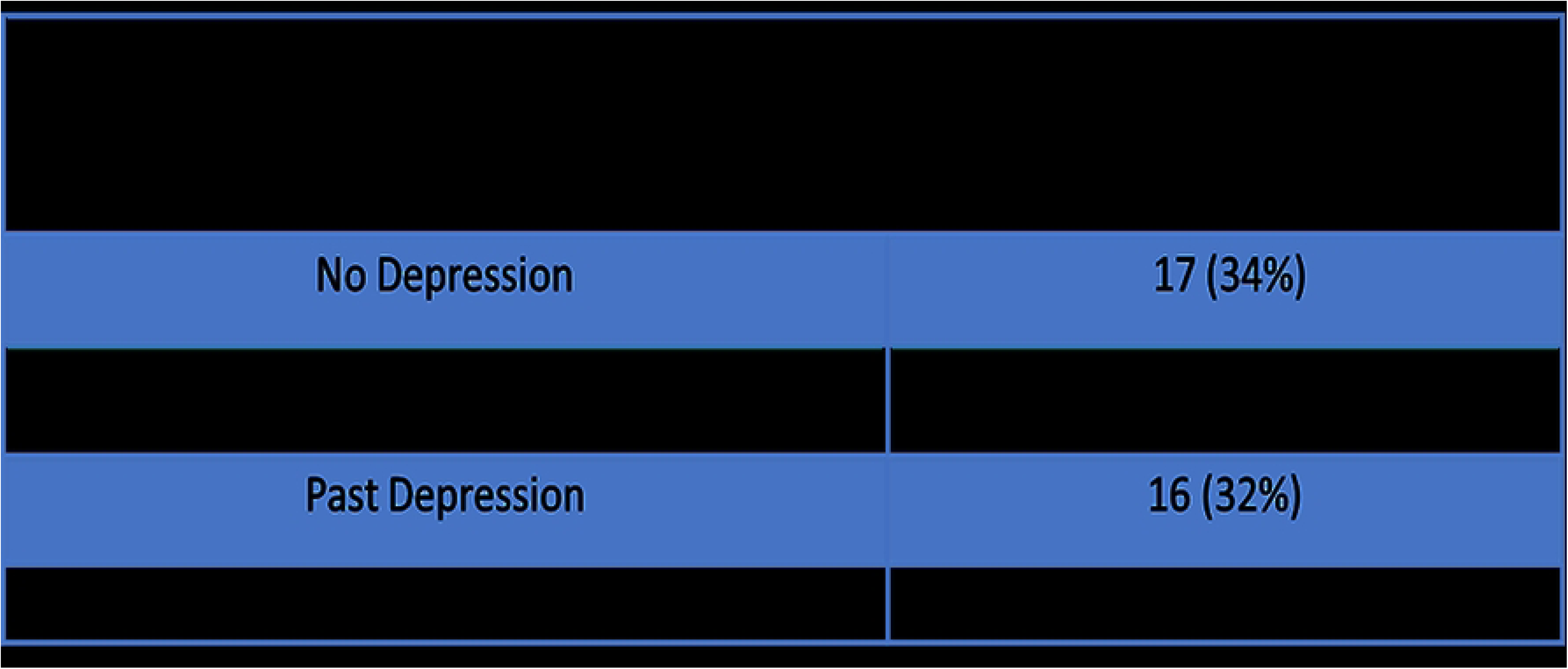
Depression confirmation by M.I.N.I. MINI outcomes of depression severity among 50 randomly selected participants from the study sample.

### Association of the socio-demographic factors with the level of depression by PHQ-9

The relation between the depression scale and the year of education has the most significant co-relation (p-value=0.000). 22 (31.4%) 1st-year medical students were found mild depressed and among them 5 (7.1%) participants severely depressed which is second-highest among all year groups. The highest portion of the second-year medical students including 20 (38.5%) participants suffers from moderate depression and this group was the biggest sufferer of very severe depression with 8 (15.4%) students. Same as the second-year medical students the third-year medical students were also populated at the moderate stage with 80 (39.8%) students. Later on, the study also found that 33 (39.8%) 4th year and 55 (45.8%) 5th year medical students suffer from moderate depression. There is a clear difference in the age gap between the students in relation to their level of depression with a significance level of 0.001. Mild to moderate depression respectively 141 (32.8%) and 151 (35.1%) students were dominant from the age group of 18-24 but 21 (4.9%) very severe cases were found among this group. On the other hand, more than half (54.6%) of the students who belong to the age group 25-29 had a moderate level of depression. In the student life money has a great role on students’ mental health (p-value=0.001), according to the survey of medical students, 12 (7.1%) students don’t have enough pocket money had a higher level of very severe depression in comparison 12 (3.4%) with those who have. And very severe depression with not getting enough pocket money has a linear relationship with no personal earnings with 19 (4.5%) students and no family contribution with 22 (4.8%) students were severely depressed. Similarly, 12 (6.6%) students having aged members in their family had a higher rate (p-value = 0.035) of very severe depression. Rather than that, caffeine consumption has a deep relation with depression (p-value=0.001). 6 (10.2%) students in the caffeine-consuming group have a prevalence of very severe depression whereas in terms of smoking 50 (52.6%) students were found with moderate depression. In comparison with males the female participants had a severity of depression with a level of significance of 0.01. 17 (5.5%) students admitted very severe depression which is only amongst 6 (2.8%) male students. The dataset shows a gradual relationship (p-value = 0.011) between depression and the level of education of the mothers of the students who participated in the study. The severity of depression decreases in comparison with the level of education of the mothers but 12 (7.9%) students were also found in the high pic of very severe depression. (Table 2).

**Table 2.**
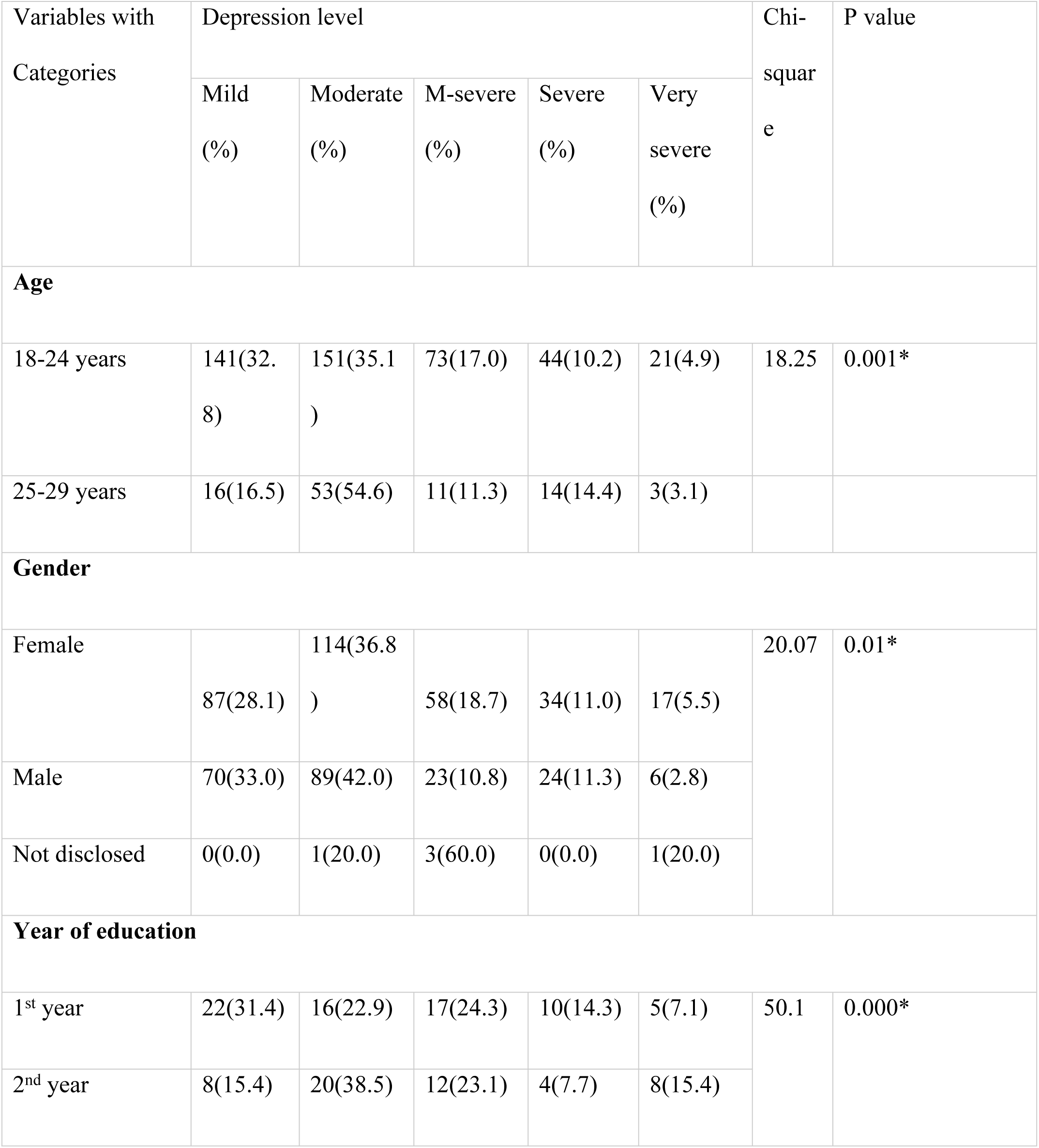
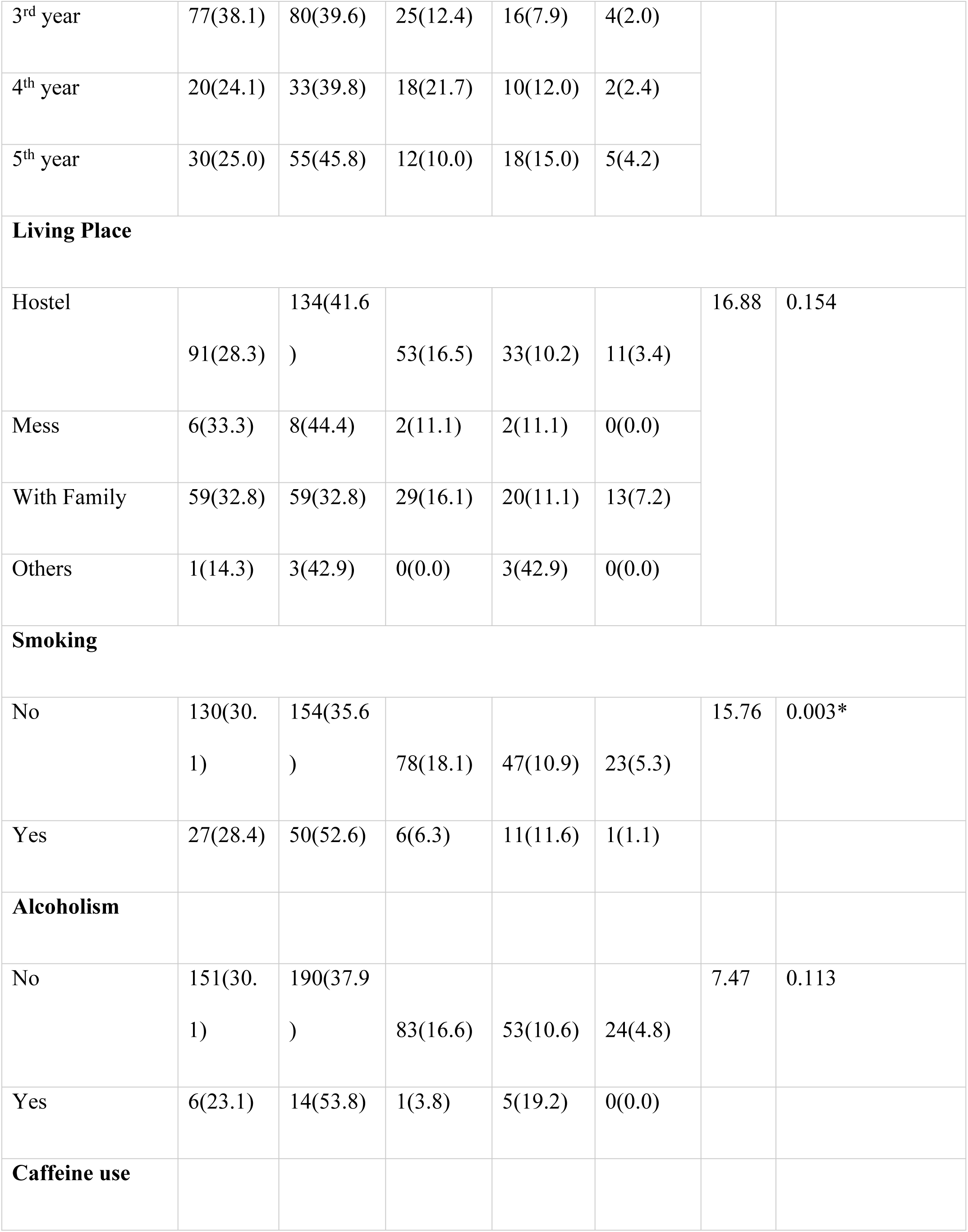
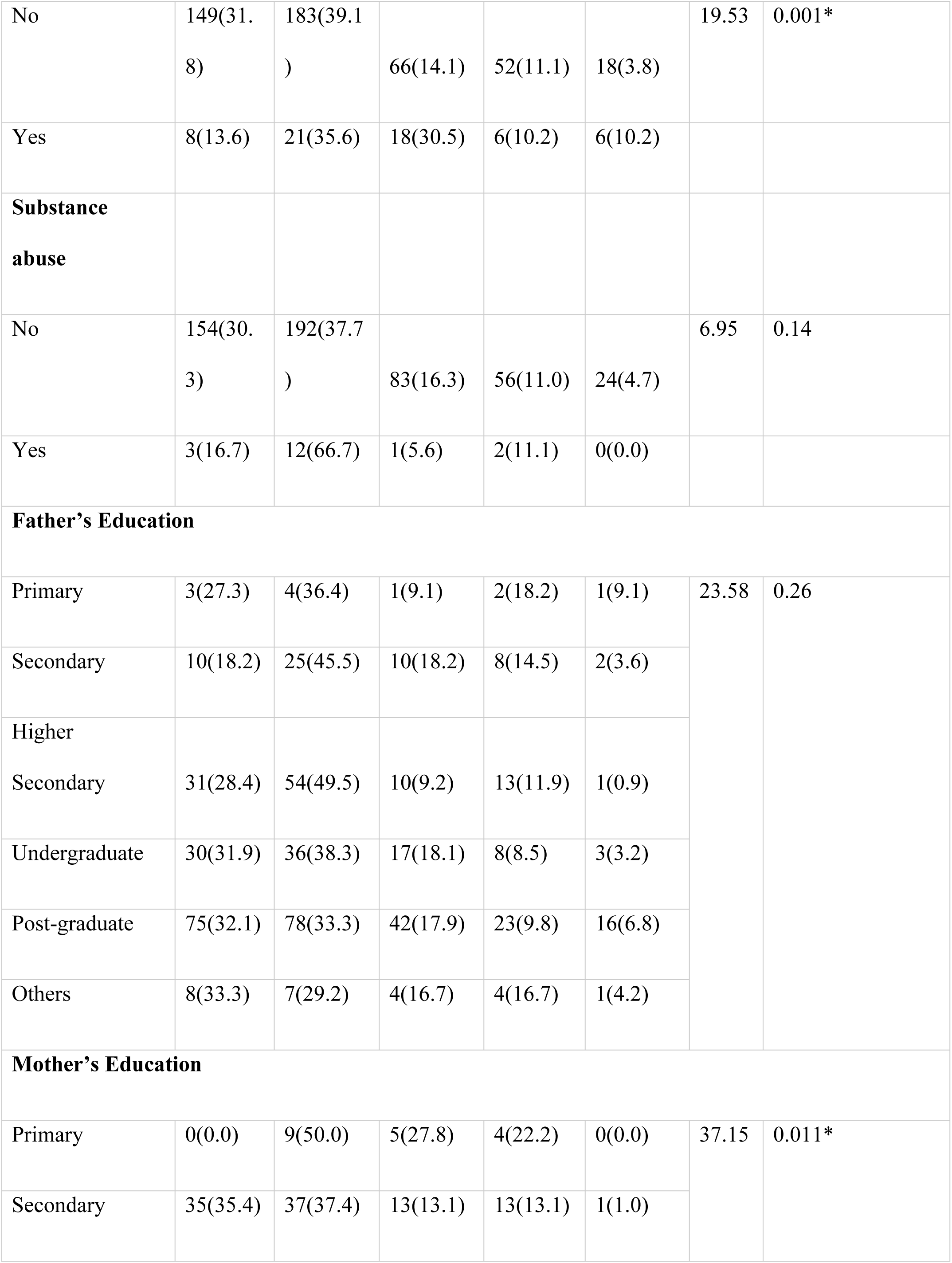
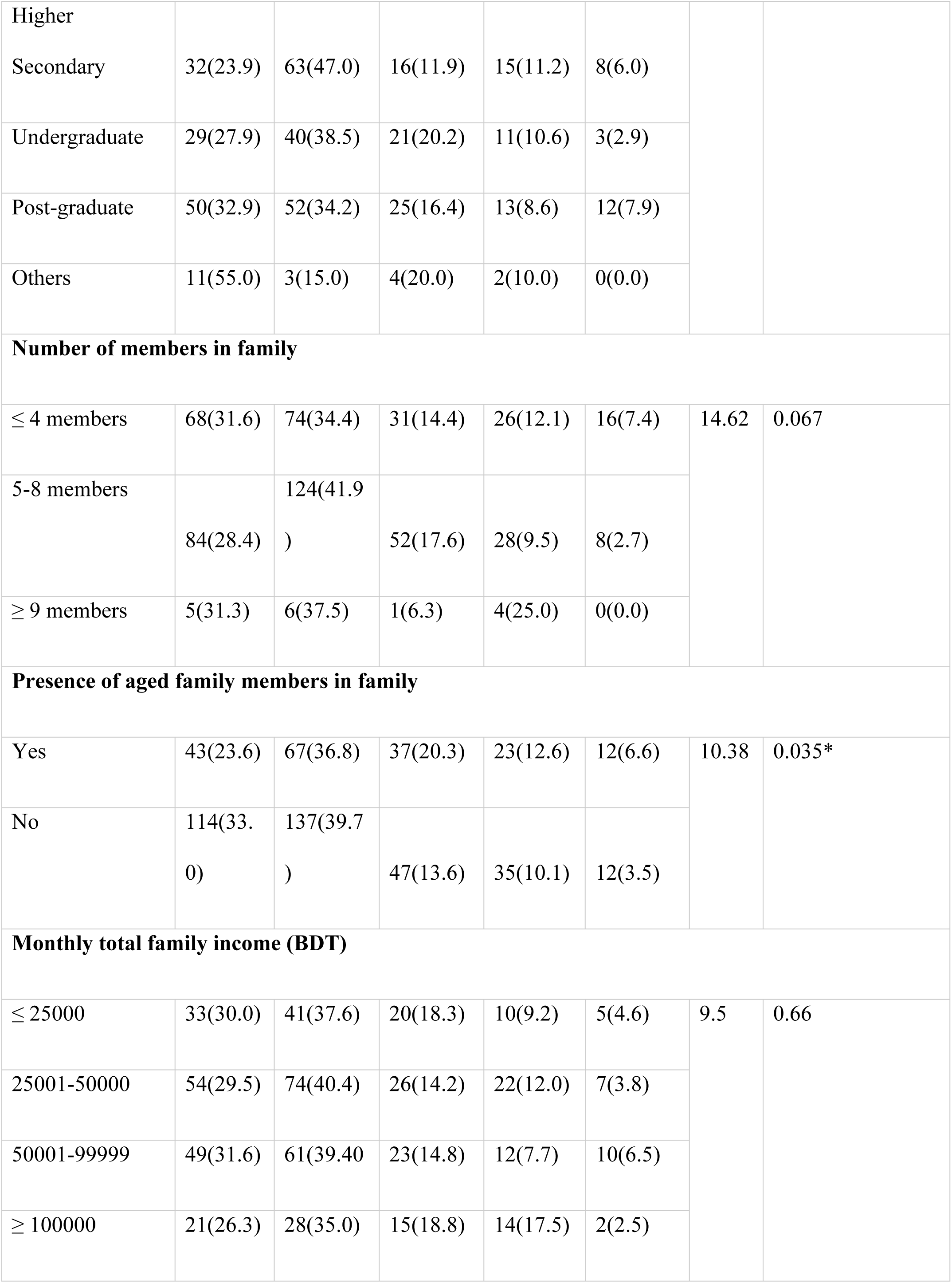
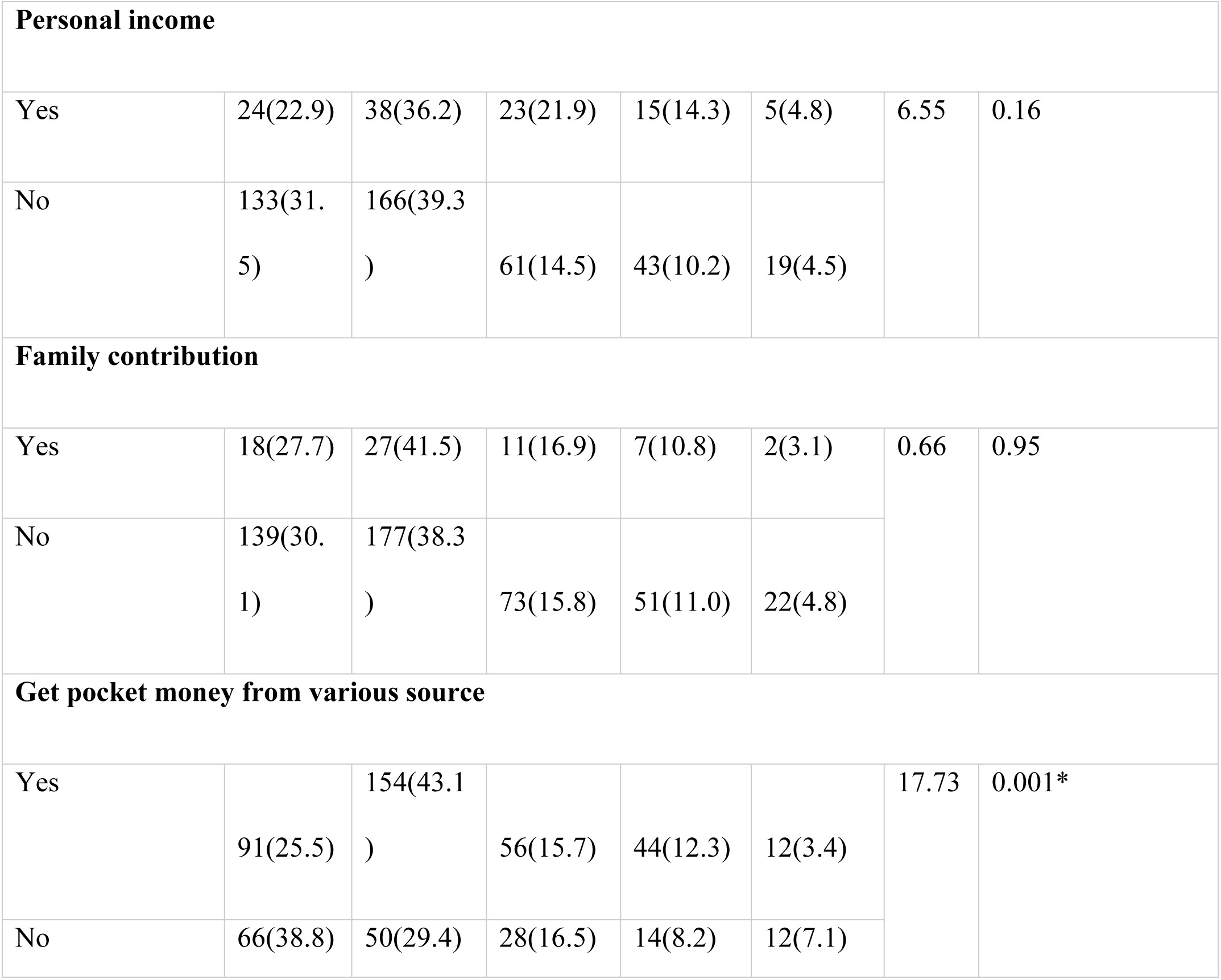
Association between sociodemographic characteristics and level of depression of study participants.

## Discussion

As the medical profession is inherently stressful, previous studies have identified doctors as one of the loneliest professional groups [29]. Depression and loneliness have been shown to be statistically significant issues among doctors [15,16]. In Bangladesh, where mental health is often stigmatized, the socio-demographic factors contributing to stress and depression are rarely acknowledged by society [32]. This study identified several factors associated with depression among medical students, including age, gender, year of education, smoking habits, caffeine consumption, maternal education, the presence of elderly family members, and receipt of pocket money. These findings align with studies conducted in South India, which also found significant associations between depression and factors such as year of study, participation in extracurricular activities, family history of depression, financial stress, substance abuse, romantic relationships, family problems, and health issues [14].

Gender differences in depression prevalence among medical students remain unclear, with some studies reporting higher prevalence among females [11,12,13], while others found it more common among males [5]. The variation in gender prevalence may depend on specific contexts and requires further exploration. Age also showed a clear correlation with depression, with younger students (aged 18–24 years) being particularly vulnerable. This study observed that newcomers to medical schools were majorly affected by depression, a finding consistent with research indicating that 60% of international students in this age group faced depressive symptoms when adjusting to Bangladeshi culture and language [31]. The relationship between gender, age, and depression warrants more in-depth investigation.

Among lifestyle-related factors, smoking was the most prevalent habit, followed by caffeine consumption, alcohol use, and substance abuse. A recent study among medical students similarly identified smoking as the leading unhealthy behavior [27]. Promoting healthy lifestyles is critical for medical students, as they are future healthcare providers and should serve as role models for their patients.

The economic background of participants’ families was relatively better compared to the national average. However, inadequate monthly allowances or pocket money were positively associated with depression [10]. This is likely due to the higher socio-economic and educational status of the participants’ parents [30]. Interestingly, the educational attainment of participants’ parents was significantly higher than the national average, suggesting a tendency for children of educated families to pursue medical careers [21]. However, the presence of severe depression in 6.8% of students highlights the potential pressure of belonging to educated families. Moreover, home-staying students appeared to be more susceptible to depression, indicating the need for institutions to organize parent-student counseling sessions to address academic and familial pressure [33].

The study also noted that over one-third of participants had elderly family members, a significant finding given that the elderly population in Bangladesh exceeds 13 million people [18]. This demographic factor may contribute to the mental health burden among students.

Approximately one-third of participants were found to have moderate to severe depression, while 38% experienced mild depression. These figures exceed those reported in previous studies using the same tools among Bangladeshi medical students [5]. The prevalence is also higher than a 2017 study conducted in Bangladesh [2]. The timing of this study, during the COVID-19 pandemic, likely exacerbated mental health issues, particularly among healthcare professionals and medical students [22]. The findings underscore the long-term vulnerability of medical students to mental health challenges, posing a significant threat to their potential contributions to future healthcare [13]. High-intensity psychosocial interventions tailored to medical students during the pandemic are urgently needed [11]. These findings align with studies conducted in Bangladesh and seven other Asian countries during the COVID-19 pandemic [2,28].

## Conclusion

This study revealed that one-third of medical students in Bangladesh experienced moderate to severe depression, while approximately 38% reported mild depression. These findings highlight an urgent need for targeted psychosocial interventions specifically designed for medical students to mitigate potential long-term impacts. Despite certain limitations, this study underscores the importance of exploring key socio-demographic factors in greater depth in future research. The results are expected to aid researchers and guide policymakers in addressing the mental health needs of Bangladeshi medical students by developing effective support systems and surveillance frameworks.

### Strengths and limitations of the study

To the best of the authors’ knowledge, the diagnostic tool MINI was used to confirm cases of depression, marking its first application among medical students in the country. Due to the COVID-19 pandemic, all interviews, including those involving the MINI, were conducted via online platforms, introducing a unique dimension to the study’s methodology. Face-to-face interviews were not feasible due to the health risks posed by the pandemic. The results should be interpreted with caution and are not generalizable, given the study’s small sample size and lack of broader representation. A key limitation was that only participants with internet access were included, restricting the study’s reach.

### Recommendations

Tackling the mental health challenges faced by medical students in Bangladesh calls for a holistic and multi-pronged strategy. Given the varied factors contributing to depression, medical institutions should focus on initiating counseling sessions, particularly targeting students who are struggling academically. Such sessions can foster a supportive environment, helping students manage academic pressures while aligning expectations with their abilities. Robust research evidence, particularly from longitudinal studies and large-scale surveys, is essential to comprehensively assess the extent of the burden. Additionally, qualitative research is needed to gain deeper insights from key stakeholders, including medical students, educators, parents, and policymakers, to better understand their perspectives and experiences.

## Data Availability

The data underlying the results presented in this study are not publicly available due to ethical and confidentiality concerns related to participants' mental health information. However, data will be made available upon reasonable request to the corresponding author. Requests must comply with ethical guidelines and institutional policies governing access to sensitive data.

## Acknowledgement

We extend our heartfelt thanks to the dedicated volunteers whose invaluable contributions to data collection were instrumental to the study’s success.

